# Smell testing to identify early alpha-synucleinopathy among people with dream enactment behavior

**DOI:** 10.1101/2024.12.11.24318857

**Authors:** Ethan Brown, Lana M Chahine, Aleksandar Videnovic, Micah J Marshall, Ryan Kurth, Chelsea Caspell-Garcia, Michael C Brumm, Christopher Coffey, Andrew Siderowf, Tanya Simuni, Kenneth Marek, Caroline M Tanner, The Parkinson’s Progression Markers Initiative

## Abstract

**Background:** REM sleep behavior disorder (RBD) is an early manifestation of alpha-synucleinopathy in many cases. Dream enactment behavior (DEB), the clinical hallmark of RBD, has many etiologies and cannot be used alone to predict underlying alpha-synucleinopathy. We compared the proportion of people with alpha-synucleinopathy, as measured by CSF alpha-synuclein seed amplification assay (CSFasynSAA), between people with polysomnographic-confirmed RBD (RBD-PSG) and people who reported DEB on a questionnaire and were further selected with smell testing and DAT-SPECT.

**Methods:** Participants were enrolled in the Parkinson’s Progression Marker Initiative (PPMI) and ≥60 years old without a diagnosis of Parkinson’s disease. Participants had either RBD-PSG or self-reported DEB. Self-reported DEB participants had to have hyposmia (<10^th^ percentile for age/sex) and at least mild DAT-SPECT abnormality (<100% age/sex-expected). We compared CSFasynSAA between RBD-PSG and self-reported DEB with hyposmia (DEB+Hypos). RBD-PSG participants also underwent smell testing and DAT-SPECT; we determined the predictive value of these tests in RBD-PSG with regards to CSFasynSAA.

**Results:** CSFasynSAA was positive in 171/240 (71%) of RBD-PSG and %) 180/210 (86%) of DEB+Hypos participants. Among RBD-PSG, hyposmia strongly predicted CSFasynSAA+ (PPV: 92% [95% CI 87%-97%]). Smell identification was more accurate than DAT-SPECT in predicting CSFasynSAA+ in RBD-PSG (AUC for UPSIT: 0.89 [95% CI 0.84 – 0.94]; AUC for DAT-SPECT: 0.65 [95% CI 0.58 – 0.73]).

**Conclusions:** Smell testing may be an effective and scalable method to identify people with alpha-synucleinopathy among those with self-reported DEB. Among individuals with RBD diagnosed by PSG, smell testing improved prediction of positive CSF alpha-synuclein biomarker.

## INTRODUCTION

Identifying individuals with early alpha-synucleinopathy is of high interest as clinical trials aiming to test interventions to reduce disability from these diseases are in the planning stages.^1^ Several non-motor and motor features may be present in early stages of alpha-synucleinopathy, but many are non-specific. Isolated REM sleep behavior disorder (iRBD) is a parasomnia defined by clinical manifestations of dream enactment behavior (DEB) and by absence of atonia during REM sleep on polysomnogram (PSG) in individuals without a diagnosis of PD or other neurodegenerative disorders.^2^ As many as 90% of people with PSG confirmed iRBD have misfolded alpha-synuclein (asyn) in their cerebrospinal fluid (CSF),^3–7^ and iRBD is now widely accepted as early-stage alpha-synucleinopathy.^8–11^ Identifying people with iRBD is a high priority to establish clinical trial-ready cohorts.^12^ ^13^

However, iRBD symptoms overlap with other disorders (e.g. sleep and psychiatric disorders) that are more prevalent and not due to neurodegeneration. The PSG required for definitive diagnosis of iRBD^14^ is not practical for large-scale screening. DEB is common – occurring in up to 19% of the general population^15^ – and can be identified in the community based on self-reported questionnaires. Yet DEB can result from other medical or psychiatric conditions and is only due to PSG confirmed RBD in a small subset.^16^ More accurate methods to identify underlying alpha-synucleinopathy in people with DEB are needed.

We therefore sought to investigate, via a tiered screening paradigm, how other assessments, specifically olfactory testing and dopamine transporter imaging, could be combined with DEB to accurately identify underlying alpha-synucleinopathy. Olfactory dysfunction is highly prevalent in neuronal alpha-synucleinopathy and is associated with disease progression in iRBD.^3^ ^17–20^ Olfactory testing has been used to identify individuals with dysfunction in dopaminergic pathways as assessed by imaging.^21^ Among people with iRBD, dopamine imaging abnormalities predict development of parkinsonism.^10^ Determining how self-report of DEB, followed by smell testing and dopamine imaging, compares to PSG in identifying individuals with iRBD would inform large-scale screening strategies that seek to identify participants eligible for clinical trials.

## METHODS

This was a cross-sectional analysis of individuals enrolled in Parkinson’s Progression Markers Initiative (PPMI), an international multisite longitudinal biomarker study.

### Sample

Participants are recruited for this study either in-clinic or through remote recruitment strategies, including media campaigns.^22^ ^23^ Inclusion and exclusion criteria are shown in Table 1. All participants had to be at least 60 years of age or older without a clinical diagnosis of Parkinson’s disease to be eligible.

**Table 1:**
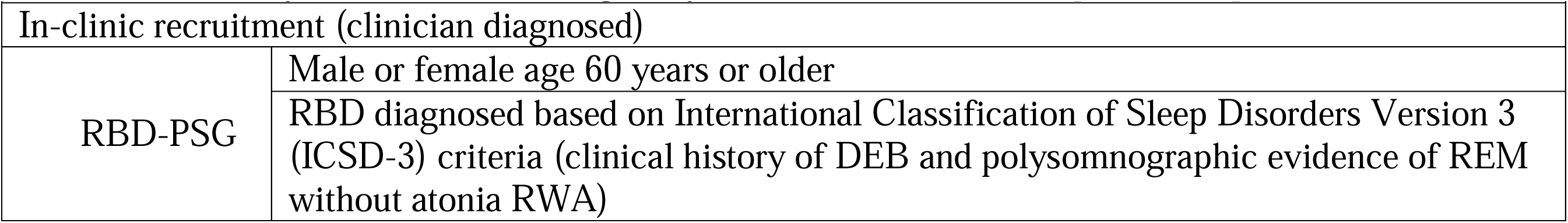
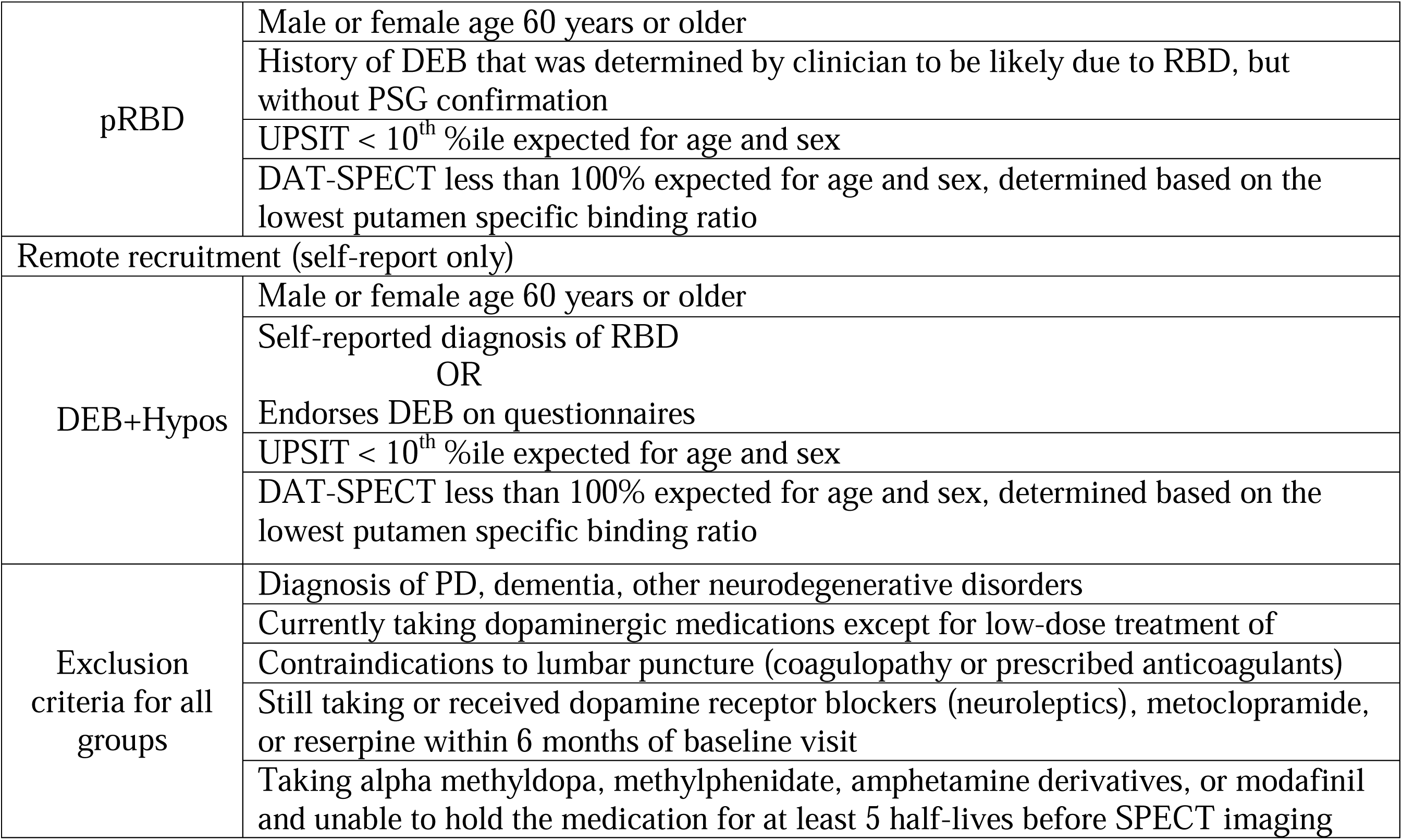
Summary of inclusion and eligibility criteria. For full criteria please see protocol.^23^.

In-clinic recruitment involved identification by study site investigators of individuals with RBD based on REM sleep without atonia on a PSG report (RBD-PSG). RBD-PSG were eligible to enroll in the study regardless of other features (iRBD). We additionally analyzed a small group of participants who were identified by investigators as having DEB thought to be likely due to RBD but without documented PSG confirmation (possible RBD, pRBD). People with pRBD also undergo smell testing with the UPSIT and dopamine transporter DAT-SPECT. People with pRBD who have an UPSIT < 10^th^ %ile and DAT-SPECT binding less than 100% expected for age and sex (determined based on the lowest putamen specific binding ratio) are eligible for enrollment.

People recruited remotely go through a staged-screening pathway previously described.^22^ Briefly, participants were identified either through PPMI Online – an online longitudinal study involving participant reported outcomes in people with and without PD^24^ – or Smell Test (ST) Direct – a series of recruitment campaigns that sent out smell tests directly to participants. Participants were included in this analysis if they completed screening questionnaires (Supplementary Table 1) and reported a prior diagnosis of RBD made by a health professional or denied a diagnosis of RBD but reported DEB based on a single item question.^25^ Participants were mailed a University of Pennsylvania Smell Identification Test (UPSIT) to complete independently online. If the UPSIT score was < 10^th^ percentile (%ile) compared to age- and sex-stratified normative values, participants were contacted for additional phone screening by a centralized team to confirm eligibility to undergo dopamine transporter imaging at a PPMI site using [123I]FP-CIT SPECT (DAT-SPECT).^26–28^ The lowest putamen specific binding ratio (SBR) was determined and compared to age- and sex-adjusted expected values. Participants with a lowest putamen SBR < 100% expected for age and sex were then invited to enroll. Because the threshold for UPSIT was much more stringent than the threshold for DAT-SPECT, where indeterminate deficits between 75% and 100% expected are less specific neurodegenerative synucleinopathy,^29^ ^30^ we refer to this group collectively as DEB+Hypos.

### Assessments

Enrolled participants underwent clinical assessments by PPMI site study personnel. Assessments included in this analysis include the Movement Disorders Society Unified Parkinson’s Disease Rating Scale (MDS-UPDRS),^31^ the REM sleep behavior disorder questionnaire (RBDSQ),^32^ and the Scales of Outcome of Parkinson’s Disease (SCOPA-Aut).^33^ Even though it was not used in selection criteria, participants with RBD-PSG also completed UPSIT and DAT-SPECT. Participants also underwent serum and cerebrospinal fluid (CSF) collection. CSF was sent for alpha-synuclein seed amplification assay (CSFasynSAA) through Amprion Inc.^6^ ^7^ ^34^ The technical document describing the assay is available for download in the PPMI database (www.ppmi-info.org/access-data-specimens/download-data). Briefly, the aSynSAA had a dual output: one for the detection or not-detection of synuclein seeds (positive, negative, inconclusive) and another for the type of seeds detected (Type 1, Type 2, and undetermined). Positive samples with Type 1 seeds presented high fluorescence values (≥45,000 RFU) and these seeds are predominantly found in participants with neuronal synuclein inclusions, while Type 2 seeds presented intermediate fluorescence values (≥3,000 RFU & <45,000 RFU) and these seeds are predominantly found in patients with underlying glial cytoplasmic inclusions and/or clinical presentation of multiple system atrophy (MSA).^35^

### Statistical analysis

We included participants who had enrolled and completed a baseline visit in PPMI with CSFasynSAA available. We first used descriptive statistics to compare the demographic characteristics and prevalence of biomarkers (UPSIT and DAT-SPECT imaging result) of participants in each group (iRBD, pRBD, and DEB+Hypos). Univariate statistics were estimated using Chi-square or Fisher’s Exact tests for categorical variables and t-tests for continuous variables. We then determined the proportion of people with positive Type 1 CSFasynSAA in each group. We separately report the proportion with positive Type 2 CSFasynSAA. Among people with RBD-PSG, we determined the association between either an UPSIT < 10^th^ %ile or a lowest putamen SBR < 75% expected for age and sex, based on the level of binding reduction observed in Parkinson’s disease,^29^ ^30^ with positive Type 1 CSFasynSAA. We then used multivariate logistic regression to determine the association between age, sex, UPSIT, and DAT-SPECT and positive Type 1 CSFasynSAA. Finally, we constructed receiver operating characteristics (ROC) curves and calculated the area under the ROC curve (AUC) to determine the accuracy of UPSIT and DAT-SPECT in identifying aSyn-SAA.

## RESULTS

### Participant selection and characteristics

From in-clinic study site recruitment, we included 240 RBD-PSG and 61 pRBD participants. Through remote recruitment, 44,865 participants at least 60 years old completed an UPSIT remotely and answered screening questions related to RBD and DEB; 1,402 (3%) endorsed an RBD diagnosis while 5,345 (12%) denied an RBD diagnosis but endorsed DEB (Figure 1). Out of these 6,747 participants, 1,907 (28%) had an UPSIT < 10^th^ %ile and 441 out of 557 (79%) who had completed DAT-SPECT had a lowest putamen SBR less than 100% expected for age and sex. While many participants are still in the process of enrolling, as of the time of data download, 210 DEB+Hypos participants had fully enrolled and had CSFasynSAA available for analysis.

**Figure 1:**
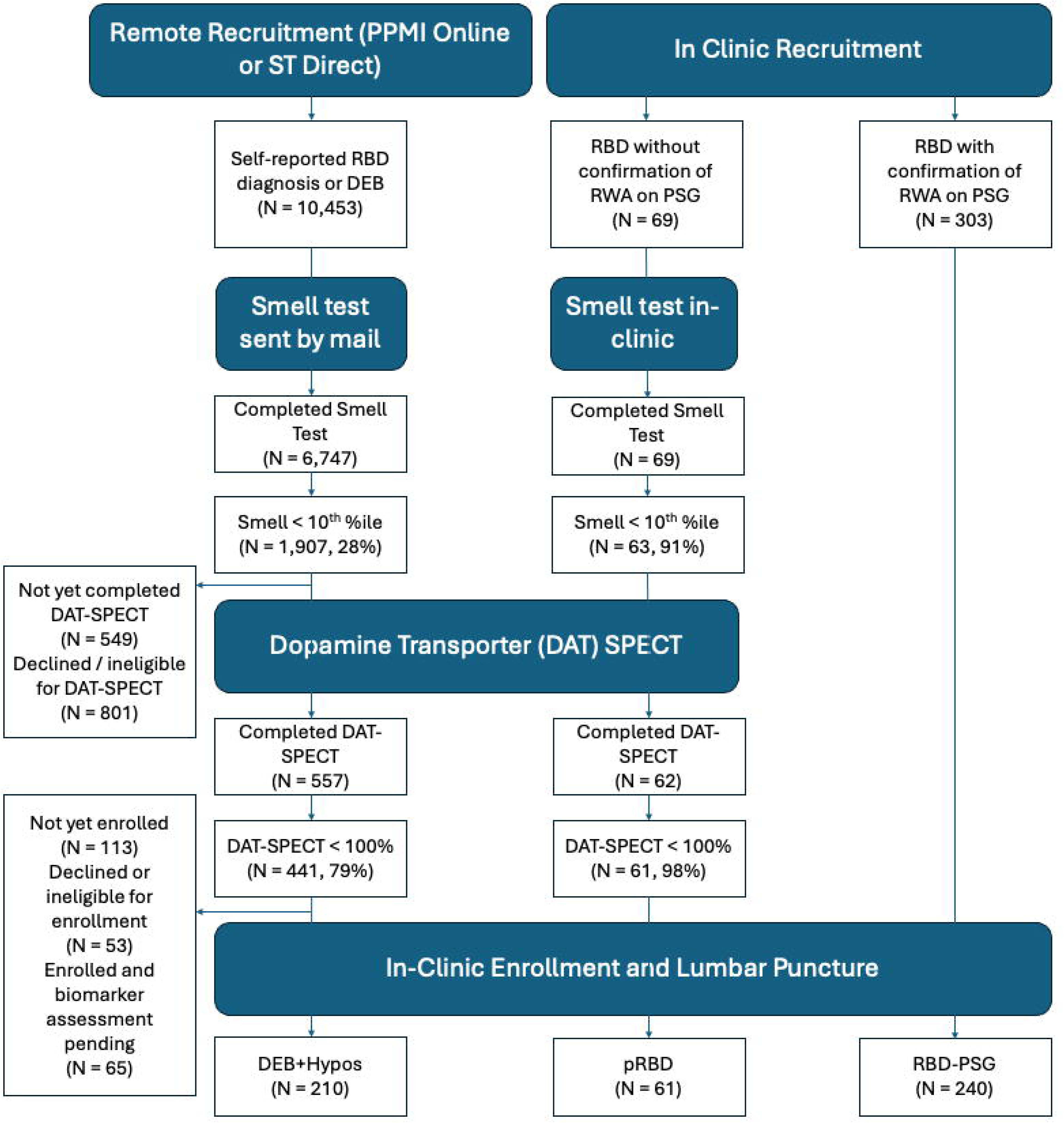
Flowchart of participants included in study. Cohorts are either recruited remotely through PPMI Online or Smell Test (ST) Direct after endorsing either a REM sleep behavior diagnosis (RBD) or dream enactment behavior (DEB), then completing a questionnaire and smell test (UPSIT) by mail (DEB+Hypos). In-Clinic recruitment involves people with RBD either with confirmation of REM sleep without atonia (RWA) on PSG (RBD-PSG) or without (pRBD).

Characteristics between RBD-PSG and DEB+Hypos are shown in Table 2; pRBD is shown in Supplementary Table 1. While age was similar in all groups, a greater proportion of the RBD-PSG group were male compared to the DEB+Hypos group.

**Table 2.** Demographic and biomarkers characteristics of participants identified either by clinical site recruitment (RBD-PSG) or by remote recruitment (DEB+Hypos).

|  | **RBD-PSG, (N=240)** | **DEB+Hypos, (N=210)** | **P-value\*** |
| --- | --- | --- | --- |
| **Age at UPSIT (years), Mean (SD)** | 67.6 (6.3) | 68.8 (5.6) | 0.1850 |
| Median (Min, Max) | 68.1 (50.6, 82.3) | 68.3 (59.6, 87.0) |  |
| **Sex, n (%)** |  |  | <.0001 |
| Female | 51 (21%) | 82 (39%) |  |
| Male | 189 (79%) | 128 (61%) |  |
| **UPSIT Percentile, Mean (SD)** | 14.7 (15.5) | 4.5 (2.4) | NA** |
| Median (Min, Max) | 8.0 (1.0, 81.0) | 4.0 (1.0, 9.0) |  |
| **UPSIT Percentile Categories), n (%)** |  |  | NA** |
| 0-10 | 139 (58%) | 210 (100%) |  |
| 10-15 | 26 (11%) | 0 |  |
| 15-100 | 75 (31%) | 0 |  |
| **DAT-SPECT % Expected, Mean (SD)** | 80.5 (25.9) | 67.5 (19.8) | NA** |
| Median (Min, Max) | 78.3 (23.6, 155.7) | 70.6 (18.6, 104.4) |  |
| **DAT-SPECT Categories, n (%)** |  |  | NA** |
| 0 - 75% | 106 (44%) | 129 (61%) |  |
| 75% - 100% | 87 (36%) | 80 (38%) |  |
| 100%+ | 47 (20%) | 1 (<1%) |  |
| **CSFasynSAA** |  |  | 0.0007 |
| Negative | 67 (28%) | 30 (14%) |  |
| Positive Type 1 | 171 (71%) | 180 (86%) |  |
| Inconclusive | 0 | 0 |  |
| Positive Type 2 | 2 (1%) | 0 |  |

\*P-values were obtained using Chi-square and Fisher's Exact tests for categorical variables and Wilcoxon Rank Sum tests for continuous variables.
\*\*Statistical test not shown as group differences are due to eligibility criteria.

### Differences in UPSIT and DAT-SPECT across groups

As expected based on inclusion criteria, the average UPSIT percentile in the DEB+Hypos group was lower than the RBD-PSG group (Table 2). DAT-SPECT was lower for the same reason, though a similar proportion had only mildly reduced DAT-SPECT binding (between 75% and 100% expected for age and sex) across groups (Table 2).

### Prevalence of CSFasynSAA positivity

Among the group recruited in-clinic, 171/240 (71%) of RBD-PSG had positive CSFasynSAA, whereas 54/61 (89%) of the pRBD group were positive. Among the remotely recruited DEB+Hypos group, 180/210 (86%) were positive (Table 2).

### Determinants of CSFasynSAA in RBD-PSG

Given the broad range of smell test results in the RBD-PSG group, we stratified the group by a smell test cutoff of the 10^th^ %ile, our inclusion criteria for other groups. Out of 240 RBD-PSG participants, 139 (58%) had an UPSIT < 10^th^ %ile. DAT-SPECT did not significantly differ between the RBD-PSG participants with and without hyposmia (Table 3). The proportion of people with positive CSFasynSAA increased: among participants with RBD-PSG and hyposmia, 92% (128/139) had Type 1 positive CSFasynSAA compared to 43% (43/101) among participants with RBD-PSG without hyposmia.

**Table 3.** Comparison of RBD-PSG participants based on UPSIT < 10th percentile (%ile).

|  | RBD-PSG ≥ 10th %ile,<br>(N=101) | RBD-PSG < 10th<br>%ile, (N=139) | P-value* |
| --- | --- | --- | --- |
| **Age at UPSIT (years), Mean (SD)** | 67.5 (5.7) | 67.7 (6.8) | 0.8317 |
| Median (Min, Max) | 68.0 (53.4, 80.3) | 68.2 (50.6, 82.3) |  |
| **Sex, n (%)** |  |  | 0.6231 |
| Female | 23 (23%) | 28 (20%) |  |
| Male | 78 (77%) | 111 (80%) |  |
| **UPSIT Percentile, Mean (SD)** | 28.2 (15.8) | 4.9 (2.4) | NA** |
| Median (Min, Max) | 25.0 (10.0, 81.0) | 5.0 (1.0, 9.5) |  |
| **UPSIT Percentile Categories), n (%)** |  |  |  |
| 0-10 | 0 | 139 (100%) |  |
| 10-15 | 26 (26%) | 0 |  |
| 15-100 | 75 (74%) | 0 |  |
| **DAT-SPECT % Expected, Mean (SD)** | 82.9 (23.7) | 78.7 (27.2) | 0.1531 |
| Median (Min, Max) | 83.3 (31.0, 152.4) | 75.7 (23.6, 155.7) |  |
| **DAT-SPECT Categories, n (%)** |  |  | 0.3257 |
| 0 - 75% | 40 (40%) | 66 (47%) |  |
| 75% - 100% | 42 (42%) | 45 (32%) |  |
| 100%+ | 19 (19%) | 28 (20%) |  |
| **CSFasynSAA** |  |  | <.0001 |
| Negative | 56 (55%) | 11 (8%) |  |
| Positive Type 1 | 43 (43%) | 128 (92%) |  |
| Inconclusive | 0 | 0 |  |

|  | RBD-PSG ≥ 10th %ile,<br>(N=101) | RBD-PSG < 10th<br>%ile, (N=139) | P-value* |
| --- | --- | --- | --- |
| Positive Type 2 | 2 (2%) | 0 |  |

\*P-values were obtained using Chi-square and Fisher's Exact tests for categorical variables and Wilcoxon Rank Sum tests for continuous variables.
\*\*Statistical test not shown as group differences are due to selection of groups.

We then determined the association between UPSIT, DAT-SPECT, and CSFasynSAA status (Figure 2; Supplementary Table 3). Among the 171 RBD-PSG participants with Type 1 positive CSFasynSAA, 128 (75%) had UPSIT < 10^th^ %ile and 85 (50%) had lowest putamen SBR < 75% expected for age and sex on DAT-SPECT. An UPSIT < 10^th^ %ile had a PPV of 92% (95% CI 88% – 97%) and 12.3 (95% CI 7.1 – 21.2, p<0.001) times increased adjusted odds of positive CSFasynSAA. DAT-SPECT < 75% expected for age and sex had a PPV of 82% (95% CI 74% - 89%) and 1.7 (95% CI 1.0 – 2.8, p = 0.05) times increased adjusted odds of having positive Type 1 CSFasynSAA (Table 4). When used as continuous measures among people with RBD-PSG to predict CSFasynSAA status, UPSIT showed an AUC of 0.89 (95% CI 0.84 – 0.94) compared to 0.65 (95% CI 0.58 – 0.73) for DAT-SPECT imaging results (Figure 3).

**Figure 2:**
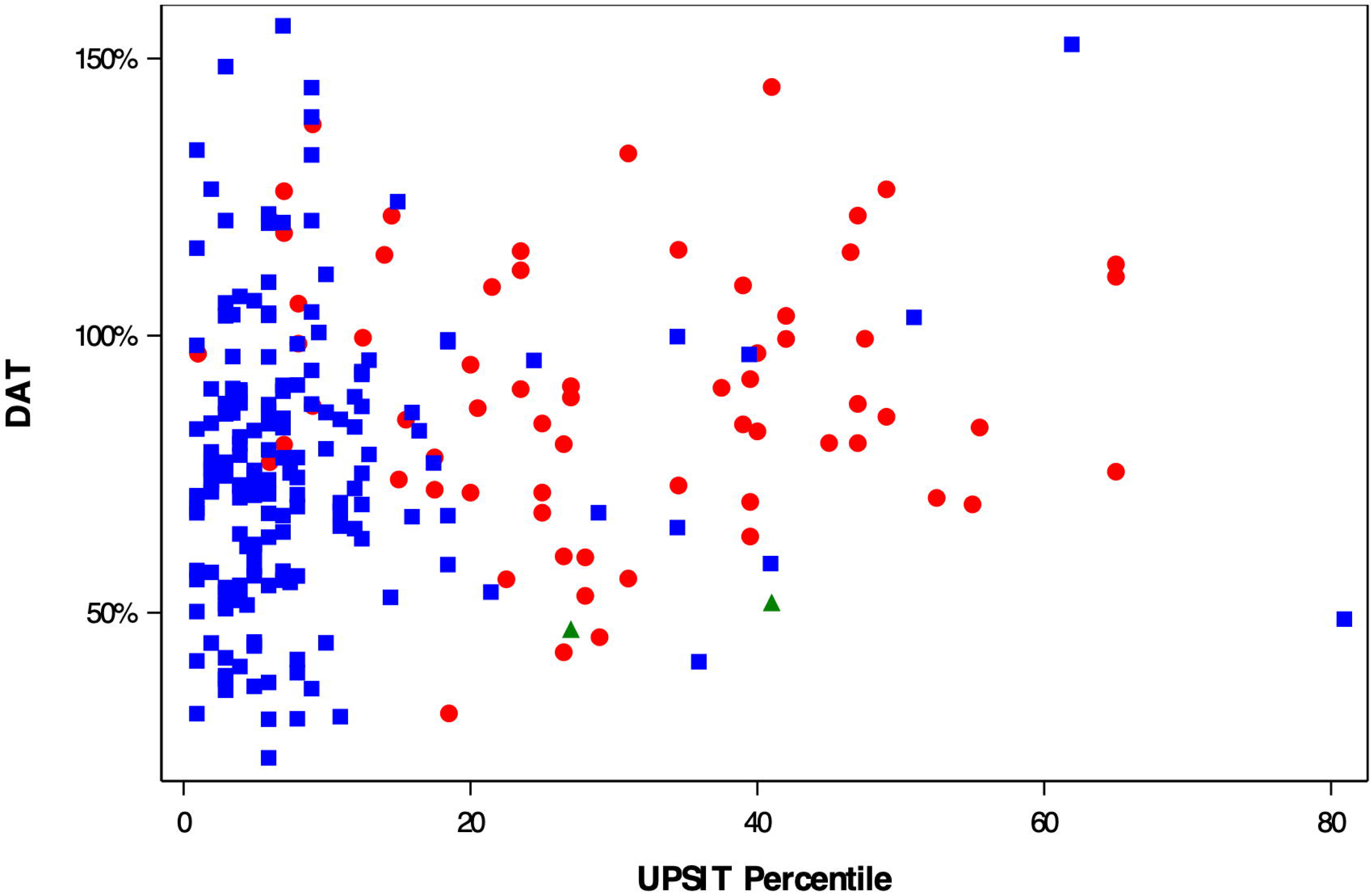
Distribution of UPSIT percentile (x-axis) compared to lowest putamen specific binding ratio percent age and sex-expected among people with RBD-PSG. Color and shape indicate CSFasynSAA results, either positive Type 1 signal (blue square), positive Type 2 signal (green triangle), or negative (red circle).

**Figure 3:**
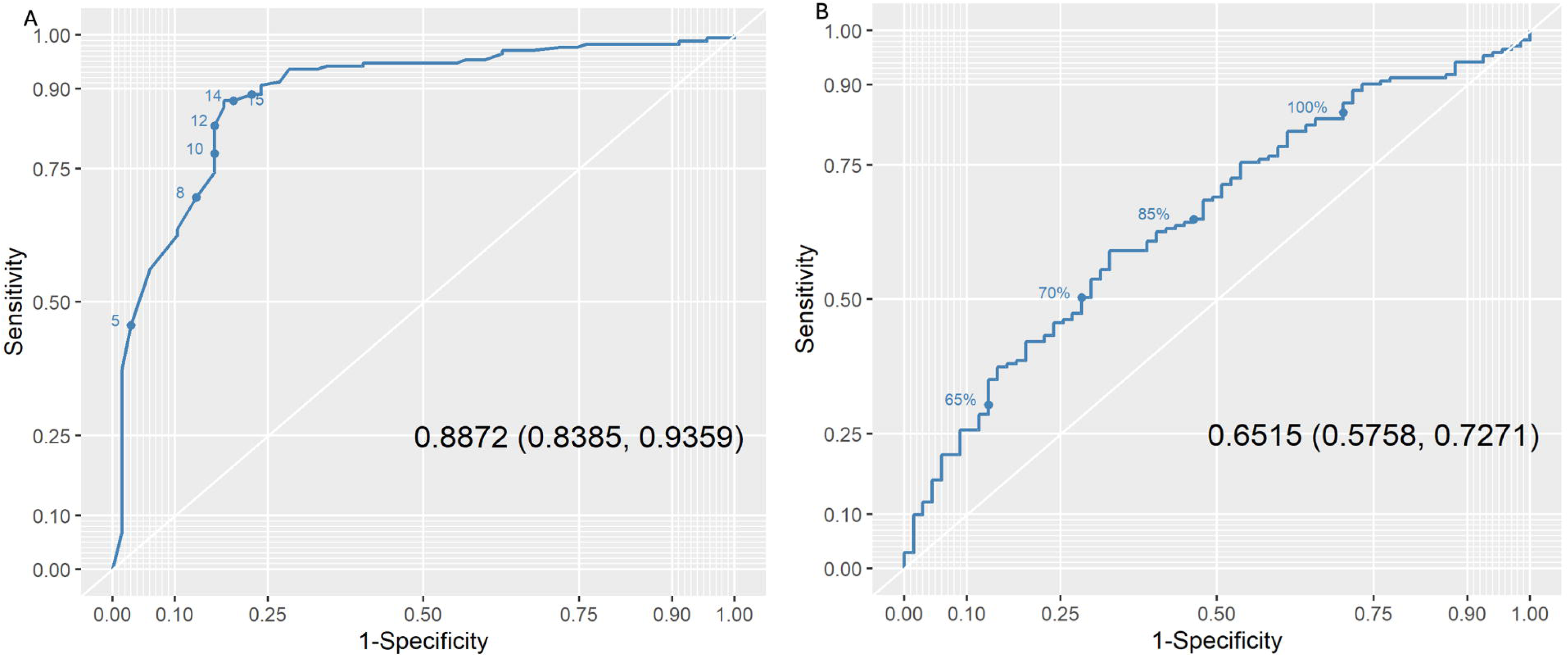
Receiver operating characteristics (ROC) curve of using an (a) UPSIT or (b) DAT- SPECT imaging results to determine who has positive CSFsynSAA among people with RBD-PSG.

**Table 4:** Multivariate regression model evaluating association between age, sex, UPSIT, DAT-SPECT, and positive Type 1 CSFasynSAA among RBD-PSG participants.

|  | Odds<br>Ratio<br>Estimate | Lower<br>CL | Upper<br>CL | p-value |
| --- | --- | --- | --- | --- |
| Age at UPSIT | 1.080 | 1.032 | 1.129 | 0.0008 |
| Sex | 2.200 | 1.266 | 3.823 | 0.0052 |
| UPSIT < 10th Percentile | 12.286 | 7.124 | 21.188 | <.0001 |
| DAT-SPECT < 75% Expected | 1.674 | 1.001 | 2.802 | 0.0496 |

### Clinical differences between participants with positive CSFasynSAA across groups

To determine whether different recruitment strategy and inclusion criteria may lead to selection for certain demographic or clinical features, we compared characteristics across participants from different groups with positive Type 1 CSFasynSAA. Demographics were similar except a higher proportion of females were seen in the DEB+Hypos group (Table 5). The MDS-UPDRS Part III was higher, though not by a large magnitude, in DEB+Hypos compared to RBD-PSG potentially due to the DAT-SPECT selection requirement. The RBDSQ was highest in the RBD-PSG with UPSIT < 10^th^ %ile group. Otherwise, no substantial clinical differences were observed. Characteristics of CSFasynSAA+ pRBD participants are shown in Supplementary Table 4.

**Table 5.** Clinical features of CSFasynSAA+ participants across groups with different recruitment strategies.

|  | RBD-PSG < 10th %ile, (N=128) | RBD-PSG ≥ 10th %ile, (N=43) | DEB+Hypos, (N=180) |
| --- | --- | --- | --- |
| Age at UPSIT (years), Mean (SD) | 68.2 (6.5) | 69.1 (4.9) | 69.0 (5.5) |
| Median (Min, Max) | 68.5 (52.7, 82.3) | 70.6 (57.3, 78.0) | 68.7 (59.6, 87.0) |
| Sex, n (%) |  |  |  |
| Female | 26 (20%) | 7 (16%) | 63 (35%) |
| Male | 102 (80%) | 36 (84%) | 117 (65%) |

|  | **RBD-PSG < 10th %ile, (N=128)** | **RBD-PSG ≥ 10th %ile, (N=43)** | **DEB+Hypos, (N=180)** |
| --- | --- | --- | --- |
| **MDS-UPDRS Total\***, Mean (SD) | 14.0 (9.9) | 14.3 (10.4) | 16.8 (10.5) |
| Median (Min, Max) | 12.0 (1.0, 53.0) | 11.5 (1.0, 57.0) | 15.0 (0.0, 53.0) |
| Missing | 2 | 1 | 5 |
| **MDS-UPDRS Part I**, Mean (SD) | 7.7 (5.6) | 7.7 (4.5) | 8.2 (5.0) |
| Median (Min, Max) | 6.0 (1.0, 27.0) | 7.0 (0.0, 17.0) | 7.0 (0.0, 26.0) |
| Missing | 0 | 0 | 1 |
| **MDS-UPDRS Part II**, Mean (SD) | 2.0 (2.9) | 2.2 (3.1) | 2.7 (3.3) |
| Median (Min, Max) | 1.0 (0.0, 16.0) | 1.0 (0.0, 12.0) | 1.5 (0.0, 18.0) |
| **MDS-UPDRS Part III**, Mean (SD) | 4.3 (4.9) | 4.3 (6.1) | 6.1 (5.9) |
| Median (Min, Max) | 3.0 (0.0, 41.0) | 3.0 (0.0, 37.0) | 5.0 (0.0, 29.0) |
| Missing | 2 | 1 | 4 |
| **RBDSQ Total**, Mean (SD) | 9.9 (2.4) | 10.4 (2.7) | 9.0 (3.0) |
| Median (Min, Max) | 10.5 (3.0, 13.0) | 11.0 (0.0, 13.0) | 10.0 (1.0, 13.0) |
| Missing | 0 | 1 | 3 |
| **SCOPA-AUT Total**, Mean (SD) | 12.7 (6.5) | 11.1 (5.4) | 12.4 (6.4) |
| Median (Min, Max) | 12.0 (0.0, 32.0) | 10.0 (2.0, 26.0) | 11.0 (2.0, 36.0) |
| Missing | 0 | 0 | 3 |
| **MoCA Total**, Mean (SD) | 26.4 (2.5) | 26.6 (2.8) | 26.7 (2.2) |
| Median (Min, Max) | 27.0 (15.0, 30.0) | 27.0 (18.0, 30.0) | 27.0 (21.0, 30.0) |
| Missing | 0 | 0 | 4 |
| **GDS**, Mean (SD) | 2.1 (2.2) | 2.4 (2.4) | 1.9 (2.0) |
| Median (Min, Max) | 1.0 (0.0, 9.0) | 2.0 (0.0, 8.0) | 1.0 (0.0, 12.0) |
| Missing | 0 | 0 | 2 |
| **STAI**, Mean (SD) | 62.9 (18.2) | 64.2 (18.7) | 59.5 (15.4) |
| Median (Min, Max) | 58.5 (40.0, 119.0) | 62.0 (40.0, 112.0) | 57.0 (40.0, 105.0) |
| Missing | 0 | 0 | 2 |

## DISCUSSION

In this study we have investigated, among individuals with dream enactment behavior, different methods of identifying early alpha-synucleinopathy. A key finding was that a questionnaire and remotely completed smell test can be used to identify a high proportion of people with positive CSFasynSAA. In our study, the proportion of people with positive CSFasynSAA was higher than in participants with RBD-PSG who were not selected based on smell. PSG remains the gold standard for diagnosing RBD but is costly^36^ and requires access to specialist sleep centers for proper administration and interpretation, which is less available in underserved populations.^37^ Questionnaires about DEB alone are not sufficient: this strategy can identify iRBD when used in people presenting to sleep clinics,^25^ but have low positive predictive value when used in the community setting.^16^ In our study, a brief questionnaire followed by a smell test and identified a population with 86% positivity for misfolded CSF alpha-synuclein. While the individuals who were identified by the questionnaire strategy could not be said to meet current diagnostic criteria for iRBD, which depends on PSG confirmation, they do have clinical and biological features consistent with early alpha-synucleinopathy. Moreover, our goal is not to make a clinical diagnosis of iRBD, but rather to identify individuals with early alpha-synucleinopathy based on positive CSFasynSAA status who are at risk for disease progression. Within this framework, the questionnaire/olfactory testing strategy is both more efficient and likely less costly than PSG.

While our algorithm included DAT-SPECT prior to SAA testing, our data further suggest that the screening protocol we used will likely be just as effective without the DAT-SPECT imaging component, which is still costly and burdensome to participants. This hypothesis is supported by our second key finding: among people with RBD-PSG, hyposmia is a strong determinant of CSFasynSAA (AUC: 0.89) compared to DAT-SPECT (AUC: 0.65). This finding is consistent with prior studies of people with RBD-PSG demonstrating more prominent hyposmia in positive compared to negative CSFasynSAA^38^ and identifying hyposmia as a risk factor for future clinical diagnosis of PD.^3^ ^10^ We therefore expect hyposmia alone to be a sufficient screen to identify early alpha-synucleinopathy among people with self-reported DEB or RBD, but future confirmatory studies are needed. Questionnaires in combination with smell testing, which can be remotely and broadly applied, would be an efficient and inclusive recruitment approach for clinical trials seeking to test therapies in people with early alpha-synucleinopathy.

Notably, the proportion of RBD-PSG participants with positive CSFasynSAA in our study (71%) was lower than what has been reported in some studies,^3^ ^6^ where cohorts of PSG-confirmed RBD have been ∼90% positive for CSFasynSAA. This difference may be because of broader inclusion in our RBD-PSG cohort compared to other studies, leading this cohort to be more heterogenous than other cohorts. In PPMI, an investigator confirms RBD based on PSG report, but full criteria may vary from center to center. Formal PSG confirmation requires stringent criteria^2^ which may improve the specificity for underlying synucleinopathy. Arguably, the criteria in PPMI are more generalizable to clinical situations and easier to implement for trials, therefore showing what could be expected in terms of enrichment for CSFasynSAA. Other recent studies have reported similar proportions of CSFasynSAA in RBD-PSG.^38^ ^39^ Importantly, even in our cohort of PSG-confirmed RBD, hyposmia significantly enriched for CSFasynSAA. UPSIT is a readily available inexpensive test that can be used broadly in sleep clinics as a proxy for likely underlying asyn pathology. However, therapeutic studies aiming to enroll iRBD participants specifically testing asyn targeting therapies will need to establish a paradigm for biomarker testing.

Our observation that DAT-SPECT was not as strongly associated with CSFasynSAA as smell supports the hypothesis that hyposmia comes prior to substantial striatal neurodegeneration. Most people with RBD-PSG and positive CSFasynSAA did not have substantial reduction in DAT-SPECT binding: ∼50% had normal or only mild reductions in DAT-SPECT binding (lowest putamen SBR ≥ 75% expected for age and sex). Longitudinal studies will test our expectation that DAT-SPECT will continue to decrease in these participants. Still, in this early stage, hyposmia appears more useful than DAT-SPECT at identifying people with underlying asyn pathology who do not yet have neurodegeneration.

Recent validation of *in vivo* alpha-synuclein biomarkers^4^ ^7^ ^40^ ^41^ has enabled a biologic definition of Neuronal Synuclein Disease (NSD),^8^ ^9^ a new terminology encompassing clinical syndromes of Parkinson’s disease (PD) and dementia with Lewy Bodies (DLB) as well as pre-diagnostic syndromes, to describe individuals with neuronal a-synucleinopathy (n-asyn). A staging system, the NSD Integrated Staging System (NSD-ISS), was also put forth, defined based on a combination of biomarkers, clinical signs and symptoms, and their functional consequences.^42^

People with biomarkers of n-asyn and possibly dopaminergic transporter (DAT) dysfunction without signs or symptoms are NSD-ISS Stage 1. Stage 2 is comprised of people with positive n-asyn only (Stage 2A), additional evidence of dopaminergic dysfunction (Stage 2B), and presence of signs or symptoms but without meaningful functional impairment. Many individuals with iRBD are in stage 2 NSD,^42^ representing a desirable cohort for intervention to prevent the onset and progression of disabling symptoms. Since n-asyn positivity is a mandatory criterion for NSD and for application of the NSD-ISS, identifying strategies that efficiently detect individuals with n-asyn positivity is crucial, and individuals with clinical history or self-report of DEB offer the prime opportunity to develop these methods. As demonstrated in this study, combining DEB with hyposmia increased the yield for identifying individuals with NSD.

### Limitations

A limitation of this study is the lack of available longitudinal data. Longitudinal data will determine how these groups may differ in the rate and type of symptoms that arise, further informing these recruitment strategies. These studies will be the focus of future analyses. Additionally, the exclusion of participants with UPSIT > 10^th^ %ile and DAT-SPECT > 100% expected in the pRBD and DEB+Hypos groups prevented us from understanding the full test characteristics of these assessments in our groups. Further information on normosmic participants with a wide range of dopaminergic function, with and without self-reported RBD/DEB, would be useful to understand these associations more completely. This selection limits the interpretation of clinical differences between these cohorts in our report. However, the goal of this work was to demonstrate the predictive value of a recruitment protocol implemented in the real world, which can be easily replicated for ongoing clinical trials targeting people with early stage NSD.

In conclusion, our study demonstrates that self-reported DEB combined with screening for severe hyposmia can identify individuals with high probability of having n-asyn pathology. This offers a scalable inexpensive approach for population screening to identify individuals in early stages of NSD. Further, assessment of sense of smell even in individuals with PSG-confirmed RBD, the current diagnostic gold standard, may be more useful than DAT-SPECT to identify people with n-asyn pathology. Smell testing could therefore provide a scalable, safe, and low-cost strategy to identify people with early stage NSD for clinical trial recruitment.

## Supporting information

PPMI Study Group Authors

## Data Availability Statement

Data used in the preparation of this article were obtained on July 8th 2024 from the Parkinson’s Progression Markers Initiative (PPMI) database (www.ppmi-info.org/access-data-specimens/download-data), RRID:SCR_006431. For up-to-date information on the study, visit www.ppmi-info.org. This analysis was conducted by the PPMI Statistics Core and used actual dates of activity for participants, a restricted data element not available to public users of PPMI data. This analysis used DAT-SPECT and aSynSAA results for participants of the Prodromal Cohort, obtained from PPMI upon request after approval by the PPMI Data Access Committee. Protocol information for The Parkinson’s Progression Markers Initiative (PPMI) Clinical - Establishing a Deeply Phenotyped PD Cohort AM 3.2. can be found on protocols.io or by following this link: https://dx.doi.org/10.17504/protocols.io.n92ldmw6ol5b/v2.

## Code Availability Statement

Statistical analysis codes used to perform the analyses in this article are shared on Zenodo (10.5281/zenodo.13951754)

## AUTHOR CONTRIBUTIONS

EB, AV, LC, AS, TS, KM, and CT contributed to the conceptualization. EB, AV, LC, CCo, AS, TS, KM, and CT contributed to the investigations. AV, LC, MD, CCo, AS, TS, KM, and CT contributed to the supervision. EB, MM, RK, CC-G, MB, and CCo contributed to the data curation and formal analysis. EB, LC, MM, RK, CC-G, MB, and CCo contributed to the validation. EB wrote the original draft. EB, AV, LC, CCo, AS, TS, KM, and CT provided project administration. EB, CCo, AS, TS, KM, and CT acquired funding. CCo, AS, TS, KM, and CT provided resources. All authors contributed to the methodology, reviewed and edited the final version, and had direct access to and verified the underlying data.

