## Supplement Tables and Figures for "Smell testing to identify early alpha-synucleinopathy among people with dream enactment behavior"

**Supplementary Table 1:** Screening questions related to REM sleep behavior disorder (RBD) and dream enactment behavior (DEB) provided to remotely recruited participants.

| Participants recruited through PPMI Online |
|---|
| 1. Do you have a diagnosis of REM behavior disorder, also known as RBD?<br>a. Yes<br>b. No<br>c. Not sure<br>d. Prefer not to answer<br><br>2. Have you ever been told, or suspected yourself, that you seem to "act out your dreams" while asleep (for example punching, flailing your arms in the air, making running movements, etc.)?<br>a. Yes<br>b. No<br>c. Not Sure<br>d. Prefer not to answer |
| Participants recruited through Smell test (ST) Direct |
| 1. Has a health care provider told you that you have a sleep problem called REM sleep behavior disorder?<br>a. Yes<br>b. No<br>c. Not sure<br>d. Prefer not to answer<br><br>2. Some people act out dreams while they sleep. They may punch, kick, yell or even fall out of bed. Have you been told you that act out your dreams? Or do you suspect you many do this?<br>a. Yes<br>b. No<br>c. Not Sure<br>d. Prefer not to answer |

**Supplementary Table 3:** Demographic and biomarkers characteristics of participants thought to have possible RBD by investigators but without polysomnography confirmation (pRBD) selected based on hyposmia and DAT-SPECT < 100% expected (see Methods).

| | **pRBD, (N=61)** |
|---|---|
| **Age at UPSIT (years),** Mean (SD) | 69.4 (5.7) |
| Median (Min, Max) | 69.3 (56.7, 81.5) |
| **Sex,** n (%) | |
| Female | 19 (31%) |
| Male | 42 (69%) |
| **UPSIT Percentile,** Mean (SD) | 6.2 (6.3) |
| Median (Min, Max) | 5.0 (1.0, 47.0) |
| **UPSIT Percentile Categories),** n (%) | |
| 0-10 | 56 (92%) |
| 10-15 | 3 (5%) |
| 15-100 | 2 (3%) |
| **DAT-SPECT % Expected,** Mean (SD) | 70.6 (21.9) |
| Median (Min, Max) | 72.3 (15.8, 111.9) |
| **DAT-SPECT Categories,** n (%) | |
| 0 - 75% | 34 (56%) |
| 75% - 100% | 26 (43%) |
| 100%+ | 1 (2%) |
| **CSFasynSAA** | |
| Negative | 5 (8%) |
| Positive Type 1 | 54 (89%) |
| Inconclusive | 1 (2%) |
| Positive Type 2 | 1 (2%) |

*P-values were obtained using Chi-square and Fisher's Exact tests for categorical variables and Wilcoxon Rank Sum tests for continuous variables.
**Statistical test not shown as group differences are due to eligibility criteria.

**Supplementary Table 3:** Test performance metrics of using either an UPSIT < 10th %ile or a lowest putamen SBR of < 75% to identify people with synSAA+ among people with iRBD.

| **UPSIT < 10th %ile** | | | | |
|---|---|---|---|---|
| Statistic | **Estimate** | **Standard Error** | **95% Confidence Limits** | |
| Sensitivity | 0.7485 | 0.0332 | 0.6835 | 0.8136 |
| Specificity | 0.8358 | 0.0453 | 0.7471 | 0.9245 |
| Positive Predictive Value | 0.9209 | 0.0229 | 0.8760 | 0.9657 |
| Negative Predictive Value | 0.5657 | 0.0498 | 0.4680 | 0.6633 |
| **DAT-SPECT < 75% expected** | | | | |
| Statistic | **Estimate** | **Standard Error** | **95% Confidence Limits** | |
| Sensitivity | 0.4971 | 0.0382 | 0.4221 | 0.5720 |
| Specificity | 0.7164 | 0.0551 | 0.6085 | 0.8243 |
| Positive Predictive Value | 0.8173 | 0.0379 | 0.7430 | 0.8916 |
| Negative Predictive Value | 0.3582 | 0.0414 | 0.2770 | 0.4394 |

| **Supplementary Table 4.** Clinical features of CSFasynSAA+ participants with pRBD. | |
|---|---|
| | **pRBD, (N=54)** |
| **Age at UPSIT (years),** Mean (SD) | 69.4 (5.5) |
| Median (Min, Max) | 69.3 (59.7, 81.5) |
| **Sex,** n (%) | |
| Female | 18 (33%) |
| Male | 36 (67%) |
| **MDS-UPDRS Total*,** Mean (SD) | 15.5 (11.3) |
| Median (Min, Max) | 13.0 (1.0, 65.0) |
| Missing | 2 |
| **MDS-UPDRS Part I,** Mean (SD) | 6.5 (5.0) |
| Median (Min, Max) | 6.0 (0.0, 21.0) |
| Missing | 0 |
| **MDS-UPDRS Part II,** Mean (SD) | 3.0 (3.7) |
| Median (Min, Max) | 2.0 (0.0, 17.0) |
| **MDS-UPDRS Part III,** Mean (SD) | 6.2 (5.8) |
| Median (Min, Max) | 5.0 (0.0, 27.0) |
| Missing | 2 |
| **RBDSQ Total,** Mean (SD) | 8.6 (3.1) |
| Median (Min, Max) | 9.5 (1.0, 13.0) |
| Missing | 0 |
| **SCOPA-AUT Total,** Mean (SD) | 12.2 (6.3) |
| Median (Min, Max) | 11.0 (2.0, 28.0) |
| Missing | 0 |
| **MoCA Total,** Mean (SD) | 25.9 (2.7) |
| Median (Min, Max) | 26.0 (19.0, 30.0) |
| Missing | 1 |
| **GDS,** Mean (SD) | 2.5 (2.9) |
| Median (Min, Max) | 1.5 (0.0, 12.0) |
| Missing | 0 |
| **STAI,** Mean (SD) | 67.7 (21.2) |
| Median (Min, Max) | 64.0 (40.0, 135.0) |
| Missing | 0 |
